# Disruptive mutations in the serotonin transporter associate serotonin dysfunction with treatment-resistant affective disorder

**DOI:** 10.1101/2023.08.29.23294386

**Authors:** Jonatan Fullerton Støier, Trine Nygaard Jørgensen, Thomas Sparsø, Henrik Berg Rasmussen, Vivek Kumar, Amy Hauck Newman, Randy D. Blakely, Thomas Werge, Ulrik Gether, Freja Herborg

## Abstract

Affective or mood disorders are a leading cause of disability worldwide. The serotonergic system has been heavily implicated in the complex etiology and serves as a therapeutic target. The serotonin transporter (SERT) is a major regulator of serotonin neurotransmission, yet the disease-relevance of impaired SERT function remains unknown. Here, we present the first identification and functional characterization of disruptive coding SERT variants found in patients with psychiatric diseases. In a unique cohort of 144 patients characterized by treatment-resistant chronic affective disorders with a lifetime history of electroconvulsive therapy, we identified two previously uncharacterized coding SERT variants: SERT-N217S and SERT-A500T. Both variants were significantly enriched in the patient cohort compared to GnomAD (SERT-N217S: OR = 151, *P* = 0.0001 and SERT-A500T: OR = 1348, *P* = 0.0022) and ethnicity-matched healthy controls (SERT-N217S: OR ≥ 17.7, *P* ≤ 0.013 and SERT-A500T: OR = ∞, *P* = 0.029). Functional investigations revealed that the mutations exert distinct perturbations to SERT function, but their overall effects converge on a partial loss-of-function molecular phenotype. Thus, the SERT-A500T variant compromises the catalytic activity, while SERT-N217S disrupts proper glycosylation of SERT with a resulting dominant-negative trafficking deficiency. Moreover, we demonstrate that the trafficking deficiency of SERT-N217S is amenable to pharmacochaperoning by noribogaine. Collectively, our findings describe the first disease-associated loss-of-function SERT variants and implicate serotonergic disturbances arising from SERT dysfunction as a risk factor for chronic affective disorders.

## INTRODUCTION

The third leading cause of disability globally is affective disorders. An estimated ∼250 million individuals have been diagnosed with unipolar depression and ∼50 million with bipolar depression, with a significant portion being treatment resistant (1, 2). A major challenge is that the underlying neurobiology of these disorders is poorly understood, a matter which is further complicated by their heterogeneous, yet partially overlapping clinical presentations (3). While bipolar and unipolar depressive disorders are defined as distinct conditions, they can both give rise to depressive episodes that are clinically indistinguishable. Moreover, both unipolar and bipolar disorders (BPD) are characterized by diverse illness trajectories, treatment responses and risk factors, including comorbid somatic and neuropsychiatric diseases (4, 5). The complex clinical presentations might reflect different or only partially overlapping disease biology as indicated by significant albeit limited genetic correlation between the disorders (6, 7). Indeed, affective disorders have complex etiologies that are still poorly understood, yet genetics is indisputably an important factor (8, 9).

Genome-wide association studies (GWAS) have found the risk for affective disorders to be highly polygenic and have identified a large number of common genetic variants with small effect sizes (10–13). Since rare variants are not captured in conventional GWASs, they could represent a significant and understudied risk factor for affective disorder for which fewer samples have been examined using exome or whole-genome sequencing techniques (14, 15). Moreover, rare variants may be helpful for deriving mechanistic insights because their potentially larger effect sizes may facilitate interrogations of their neuropathological impact.

The serotonergic system of the brain has been convincingly implicated both in pathophysiological processes of affective disorders and in determining treatment responses (16–18). Indeed, first-line treatment for depression is selective 5-HT reuptake inhibitors (SSRIs) targeting the serotonin transporter (SERT), which is localized in the presynaptic membrane of serotonergic neurons. By mediating serotonin (5-HT) reuptake, SERT exerts tight control over extracellular 5-HT dynamics and ensures both termination of 5-HT signaling and replenishment of the intracellular neurotransmitter pool (19). SERT is therefore critical for fine-tuning the 5-HT signals that ultimately encode its modulatory functions on memory, learning and reward processes and influences emotional states linked to mood, anxiety and aggression (17, 20). Accordingly, 5-HT-regulated neuronal processes and behaviors can be affected by changes in SERT function.

Several observations support the hypothesis that impairments in SERT function could be a risk factor for affective disorders (20–26). For instance, blocking 5-HT reuptake during early development in rodents causes depression-related phenotypes later in adulthood (22, 23). Additionally, positron emission tomography studies have found a pattern of low midbrain SERT levels in patients with MDD (24) whereas the level of SERT increases in patients recovering from a depressive episode (25). However, the etiological link between SERT and affective disorder has not been consolidated by GWAS and remains unclear. Compared to studies on common variants, the possible role of rare coding SERT variants in affective disorders is largely unexplored. Importantly, no loss-of-function coding variants of SERT have yet been linked to disease, while two rare gain-of-function coding variants of SERT, SERT-G56A and SERT-I425V (27–29), have been reported to associate with Autism Spectrum Disorder (ASD) and Obsessive Compulsive Disorder (OCD), respectively (27, 30).

Here, we provide new insight into the role of rare coding variants of SERT in affective disorders. In a unique cohort (31) of 144 ethnic Danish treatment-refractory uni-and bipolar depression patients with a lifetime history of electroconvulsive therapy (ECT), we identify two coding SERT missense mutations, SERT-N217S and SERT-A500T, in three unrelated patients. Both variants were found at a significant higher frequency in the ECT cohort compared to control populations. Our study also reveals that SERT-N217S and SERT-A500T share a ‘loss-of-function’ molecular phenotype and that the functional impairment in SERT-N217S arises from disrupted glycosylation, which in turn drives impairments in trafficking and surface expression. Finally, we show that SERT-N217S also exerts dominant-negative effects on wild-type SERT (SERT-WT) and that pharmacological chaperones can be applied to rescue SERT-N217S function.

Collectively, our investigation identified the first hypomorphic mutations of SERT in patients with psychiatric disease. The observations support that serotonergic dysfunction can be a risk factor for treatment-resistant affective disorders.

## METHODS AND MATERIALS

### Subjects: genetic and diagnostic information

SERT coding variants were identified in a whole exome sequence dataset from a previously described Danish cohort of treatment-refractory uni-and bipolar ECT patients (31). Exome sequencing of DNA from blood samples was done using the Illumina Nextera Rapid Capture Kit at an average of 20x depth. The sequence reads were aligned to the human reference genome hg19 using the Burrows-Wheeler short read aligner (32). The MarkDuplicates tool (https://broadinstitute.github.io/picard/) was used to identify duplicate reads and variant discovery was carried out using HaplotypeCaller (33). Out of 144 ethnic Danish patients from the cohort, 10 (6.9%) were carriers of a SERT missense mutation, all being heterozygote, and this included 7 (4.9%) carrying SERT-G56A, 2 (1.4%) carrying SERT-N217S and 1 (0.7%) carrying SERT-A500T. Sanger sequencing was used to verify the presence of the SERT variants. The kinship coefficients for the 144 samples were calculated using the relationship inference algorithm KING, which found no familial relationships among the samples (34).

Information about the occurrence of SERT-G56A, SERT-N217S and SERT-A500T in ethnicity matched controls were provided by look up in an exome sequenced control sample from the iPSYCH consortium’s first phase genotyping of a nation-wide Danish birth cohort (35), following sample and variant quality control, performed as described in (36).

Diagnostic information for each patient and their first-degree relatives was obtained from the Danish nationwide registers on health issues.

### Cell Culturing and Transfection

HEK293 cells were grown in DMEM with 10% dialyzed FBS and 100µg/mL penicillin-streptomycin and maintained and transfected as described previously (37, 38). All experiments were carried out in cells transiently transfected with WT-SERT, SERT-N217S and/or SERT-A500T in pcDNA3 expression vector using Lipofectamine 2000 (Invitrogen) and assays were conducted 36-48 h after transfection. The N217S and A500T mutations were generated on WT-SERT background using a QuikChange site-directed mutagenesis kit (Stratagene) and verified by sequencing.

### Co-localization Assay

For the co-localization assay, transiently transfected HEK293 cells were seeded on poly-L-ornithine-coated coverslips. The following day, the cells were washed in ice-cold PBS before fixating them with 4% paraformaldehyde in PBS on ice for 10 min followed by 10 min at room temperature. The cells were washed twice with blocking buffer (1% BSA in PBS) and incubated in blocking buffer for 20 min. After that, the slides were incubated with primary antibodies in dilution buffer (0.2% saponin in PBS) for 1 h before washing them three times in blocking buffer and then incubating them for 1 h with secondary antibodies in dilution buffer. Finally, the coverslips were washed once in blocking buffer, twice in PBS and once in Milli-Q water before mounting them with ProLong Gold Antifade Mountant. Antibodies were used at the specified dilutions:

Goat anti-SERT (Santa Cruz Biotechnology, sc-1458; discontinued, 1:100), rabbit anti-Giantin (Covance, PRB-114C, 1:800), rabbit anti-GRP78/BiP (Abcam, AB21685, 1:200), rabbit anti-Lamp1 (Abcam, AB24170, 1:800), Alexa Fluor 488 conjugated donkey anti-goat (Invitrogen, A11055, 1:500), Alexa Fluor 568 conjugated donkey anti-rabbit (Invitrogen, A10042, 1:500).

### SERT-labeling with VK02-83

The day before labeling, transfected HEK293 cells were seeded on poly-L-ornithine-coated Lab-Tek II 4-well Chambered Coverglass (Nunc). For labeling, the cells were incubated with the highly SERT-selective fluorophore, VK02-83 (40nM) (39) for 30 min at 4 °C following three washes with ice-cold PBS and live imaging was carried out immediately afterward. For negative control, WT-SERT expressing cells were incubated with 1 µM paroxetine for 15 min before co-incubation with VK02-83.

### [^3^H]-5-HT Uptake

[^3^H]-5-HT saturation uptake curves were obtained as described previously (40). See Supplementary Information for detailed experimental protocol and analysis.

### Imaging

Confocal imaging was carried out with a Zeiss LSM 510 laser-scanning microscope using an oil-immersion plan-apochromat 63x/1.4 DIC objective. Alexa Fluor 488-conjugated antibody was excited with a 488nm argon laser followed by detection of emitted light with a 505-550nm band-pass filter. Both Alexa Fluor 568-conjugated antibody and VK02-83 were excited with a 543nm helium-neon laser followed by detection of emitted light with a 560nm long-pass filter. The quantification of VK02-83 membrane staining and the Pearson correlation coefficient for co-localization were performed in ImageJ using macros. See Supplementary Information for detailed protocol and macros used.

### Surface biotinylation and Western blotting

Surface biotinylation and Western blotting were performed as previously described (37) using primary goat anti-SERT antibody (Santa Cruz Biotechnology, sc-1458; discontinued, 1:750) and secondary horseradish peroxidase-coupled rabbit anti-goat antibody (Pierce, 1:5000). Protocol is detailed in Supplementary. PNGase F treatment of samples were performed according to the manufacturer’s protocol (New England Biolabs). For rescue experiments, the cells were treated with or without noribogaine (10 µM) for 24 h before performing surface biotinylation and western blotting.

### Study approval

Index patients were included from a previously described Danish cohort of ECT treated patients, which was approved by the Danish Ethical committee and Data Protection Agency and with written informed consent obtained from patients (31). The Danish Scientific Ethics Committee, the Danish Data Protection Agency and the Danish Neonatal Screening Biobank Steering Committee approved the iPSYCH study (41). Genetic information from the iPSYCH cohort (35) does not require informed consent as it classifies as register-based research.

## RESULTS

### Identification of SERT coding variants in treatment-resistant patients with affective disorders

To address the hypothesis that rare functional SERT variants may be a risk factor for affective disorder, we identified all coding variants in the *SLC6A4* gene in a unique sample of therapy-resistant patients suffering from a chronic affective disorder. This cohort has been described previously (31) and is comprised of Danish patients, who were diagnosed with chronic affective disorder (i.e. bipolar and unipolar major depressive disorder) based on the criteria of International Classification of Diseases (ICD)-10 and who all had a lifetime history of ECT According to current treatment guidelines, ECT is used only for severe and treatment-resistant forms of affective disorders (42).

From the cohort, we obtained exome-sequencing data from these patients and identified three coding SERT variants among 10 (6.9%) unrelated heterozygote carriers. These included 7 carriers of SERT-G56A (4.9%), 2 carriers of SERT-N217S (1.4%), and 1 carrier of SERT-A500T (0.7%) (Table 1). Subsequent Sanger sequencing confirmed the genotypes. The most prevalent coding SERT variant (41), SERT-G56A, has already been extensively studied and demonstrated to be a gain of function variant (28, 43, 44). By contrast, we could not find any characterization of either SERT-N217S or SERT-A500T in the literature.

**Table 1.** The two SERT coding variants, SERT-N217S and SERT-A500T, are significantly enriched in the ECT cohort. The number of alleles (n) with the SERT-G56A, SERT-N217S and SERT-A500T mutations, and the overall number of alleles in the ECT cohort, the GnomAD database, and in an ethically matched group of healthy controls from the iPSYCH2012 cohort (41). Association analysis was performed for each SERT variant in the ECT cohort to derive the odds ratios (OR) with 95% confidence interval (CI) relative to the control populations, GnomAD and iPSYCH. Fisher’s exact tests were used for the statistic. Symbols: *, P < 0.05; **, P ≤ 0.01; ***, P ≤ 0.001.

| SERT variant | Number of alleles (n) |  |  |  |  |  |  |  |  |
| --- | --- | --- | --- | --- | --- | --- | --- | --- | --- |
|  | ECT cohort<br>n | GnomAD |  |  |  | iPSYCH |  |  |  |
|  |  | n | P | OR | 95% CI | n | P | OR | 95% CI |
| G56A<br>(c.167C>G) | 7 | 2495 | ns | 1.27 | 0.60 - 2.68 | 243 | ns | 0.98 | 0.46 - 2.04 |
| N217S<br>(c.650T>C) | 2 | 6 | *** | 151 | 30.3 - 749 | $\leq 4^{\#}$ | * | $\geq 17.7$ | $\geq 3.75 - 83.2$ |
| A500T<br>(c.1498G>A) | 1 | 0 | ** | 1348 | 54.8 - 33160 | 0 | * | ∞ | $3.77 - \infty$ |
| Total allele number | 288 | 129186 |  |  |  | 9108 |  |  |  |

<sup>#</sup>GDPR rules prevent reporting on fewer than 5 individuals from the iPSYCH cohort.
Accordingly, in those cases, the n, OR and 95%CI values are reported for $n \leq 4$ .

To investigate if each of the identified coding variants accumulated in the patient cohort, we compared their allele frequencies with that found for Europeans in the Genome Aggregation Database (GnomAD) (45) (v2.1.1) (Table 1). In this database, the SERT-G56A variant had a reported allele frequency of 0.01931 (2495/129204), SERT-N271S had a frequency of 0.00004644 (6/129186) while SERT-A500T was not observed (Table 1). Of notice, the GnomAD database does not contain information on clinical diagnosis. Hence, we cannot tell if the carriers found in this database have relevant psychiatric diseases. Nevertheless, we found a significant accumulation of SERT-N217S (OR = 151) and SERT-A500T (OR = 1348) in our ECT cohort compared to GnomAD (Table 1). In contrast, no enrichment was observed for SERT-G56A (OR = 1.27, Table 1). We also used available exome sequence data from 4885 Danes known *not* to hold a diagnosis of attention-deficit/hyperactivity disorder, autism spectrum disorder, bipolar, schizophrenia, and single or recurrent depression, to evaluate the occurrence of the SERT variants among ethnically-matched healthy controls subjects. Compared to this control population, which is part of the IPSYCH2012 cohort (41), we again found that the SERT-N217S and SERT-A500T variants were enriched in the ECT cohort, (SERT-N217S: OR ≥ 17.7 and SERT-A500T: OR = ∞, Table 1), while no difference in allele frequency was observed for SERT-G56A (OR = 0.98, Table 1)

Collectively our genetic data suggest that SERT-N217S and SERT-A500T could be functional variants, while SERT-G56A does not appear to increase risk for affective disorder. Since SERT-G56A has also been functionally characterized (28, 43, 44), we focused the remainder of our investigations on the disease-associated and uncharacterized SERT-N217S and SERT-A500T variants.

Unfortunately, none of the patients were available for further studies, nor were relatives. However, records of psychiatric diagnoses were available for the patients and first-degree relatives via the Danish health registers. The register data contains records of all patients’ contacts with Danish non-psychiatric hospitals since 1977 and psychiatric hospitals since 1995. Each contact includes one primary and optional secondary ICD diagnosis code (46). Figure 1A shows pedigrees containing diagnostic information for each patient and their first-degree relatives. The first patient carrying the SERT-N217S variant is deceased (Figure 1A, left). She was diagnosed with neurasthenia, depressive episodes, dissociative motor disorders, anxiety disorder, adjustment disorder and bipolar disorder. Her three sons have all been diagnosed with alcohol use disorder, two of them with anxiety disorder and two with unspecified reactions to severe stress. Additionally, one had a registered suicide attempt and one had experienced recurrent depressive episodes. The children’s father did not have any psychiatric diagnoses registered.

**Figure 1.**
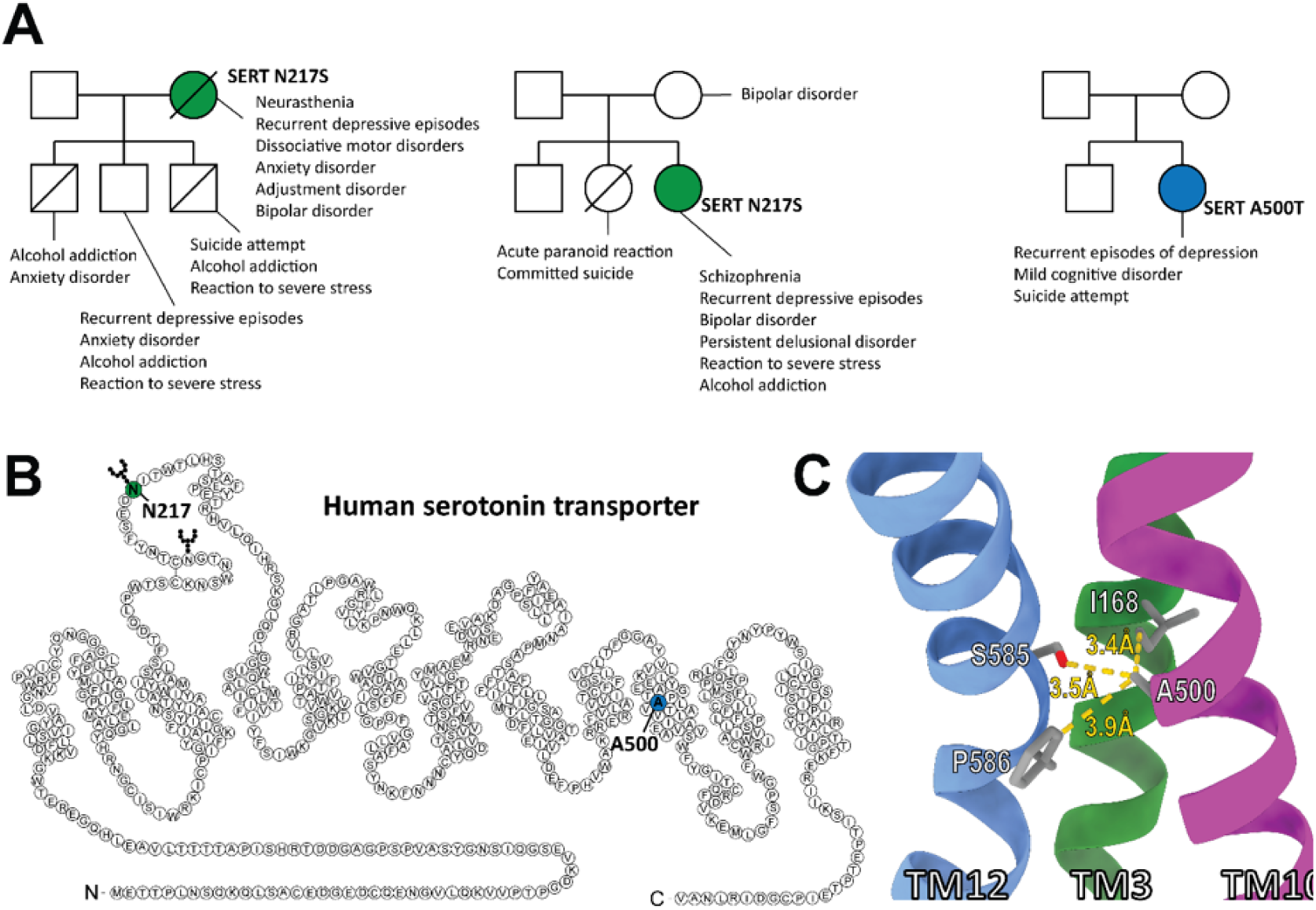
Identification of two uncharacterized SERT coding variants in patients with refractory affective disorder. **(A)** Pedigrees of three unrelated families with probands (colored) with chronic affective disorder and carrying a SLC6A4 coding variant. Two probands carry the same N217S mutation and one carries an A500T mutation. All psychiatric diagnoses received, according to the Danish National Patient Registry, are included for each family member. Symbols: Square = male; circle = female; slash = deceased. **(B)** Snake diagram of the human SERT with the location of the two missense mutations annotated. The SERT-N217S variant affects one of two glycosylation sites of SERT in extracellular loop 2, while the SERT-A500T mutation is located in the center of TM10. **(C)** Three-dimensional model of the interface between TM3, TM10 and TM12 of human SERT where A500 is located. Residues within 4 Å of A500 are shown.

The second patient carrying SERT-N217S was diagnosed with schizophrenia, bipolar disorder, persistent delusional disorder, unspecified stress reaction and alcohol addiction. Her mother was diagnosed with bipolar disorder and her sister was diagnosed with acute paranoid reaction and committed suicide in her late teens. Neither her father nor brother had any psychiatric diagnoses.

Finally, the patient carrying SERT-A500T had diagnoses of recurrent episodes of depression and mild cognitive disorder and a record of attempted suicide. None of the close relatives of the SERT-A500T carrier had psychiatric diagnoses.

Collectively, although we do not know the genetic background for the relatives of the carriers, we observed an apparent overrepresentation of affective phenotypes in the relatives of the two SERT-N217S carriers.

### SERT-N217S and SERT-A500T are both loss-of-function variants

Figure 1B illustrates a snake diagram of SERT with annotation of the SERT-N217S and SERT-A500T missense mutations. The asparagine at position 217 is the second of two glycosylation sites in SERT located in extracellular loop 2 (47, 48). The predictive Polyphen-2 (49) and SIFT (50) tools score the N271S mutation as benign and tolerated, respectively. In contrast, the SERT-A500T mutation is scored as possibly damaging and deleterious where residue position 500 is located in the center of transmembrane helix 10 (TM10). Zooming in on human SERT A500 (PDB ID: 6AWO (51), Figure 1C) shows that this residue is situated in a tightly spaced region, facing the interface between TM3, TM10 and TM12. From the β-carbon of A500, the distance to both p.I168 in TM3, and p.S585 and p.F586 in TM12 are around 3-4 Å. Thus, it is plausible that a mutation from alanine to the bulkier threonine could distort the transmembrane domain.

To directly investigate the functional impact of the SERT-N217S and SERT-A500T mutations on 5-HT uptake kinetics, we performed homologous competitive uptake experiments on HEK293 cells transiently transfected with either SERT-WT, SERT-N217S, or SERT-A500T (Figure 2A). Remarkably, we found that both variants show functional impairments. Thus, for SERT-N217S we observed, in contrast to what was predicted by Polyphen-2 (49) and SIFT (50), a pronounced reduction in uptake capacity compared to SERT-WT of 54.2 ± 6.2% (*P* < 0.0001, Figure 2A) accompanied by a slightly higher apparent 5-HT affinity (*K*_m_ = 239 ± 70 nM) than SERT-WT (*K*_m_ = 655 ± 106 nM, *P* = 0.016, Figure 2A). The SERT-A500T variant, which was predicted damaging by *in silico* analysis, also showed a reduction in uptake capacity of 36.9 ± 4.5% (*P* = 0.0002, Figure 2A), but without changes in apparent 5-HT affinity (*K*_m_ = 966 ± 70 nM, Figure 2A) compared to SERT-WT.

**Figure 2.**
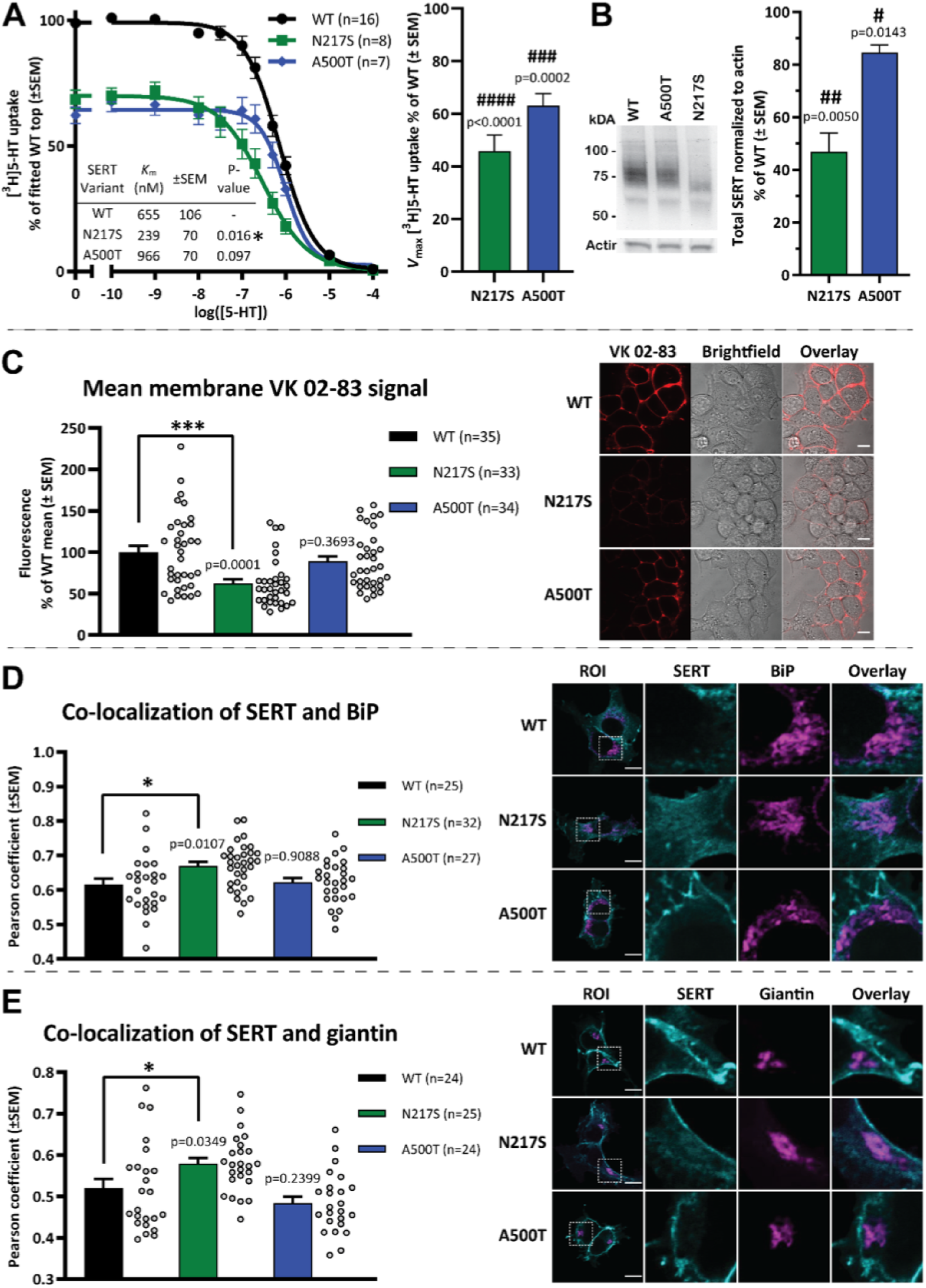
Kinetic properties and cellular processing of SERT-N217S and SERT-A500T. **(A)** (Left) Homologous competitive uptake curves from transiently transfected HEK293 cells with apparent 5-HT affinities, and *V*_max_ values (right). Both SERT-N217S and SERT-A500T demonstrated a reduced maximum transport rate of 5-HT transport and SERT-N217S displayed a significantly higher apparent 5-HT affinity than WT-SERT. *K*_m_ and *V*_max_ values were determined by fitting the uptake curves to Michaelis-Menten kinetics. The *V*_max_ values were, for each experiment, calculated as % of SERT-WT, which had a mean absolute *V*_max_ of 3246 ± 498.5 fmol/min/10^5^ cells. All experiments were performed in triplicates with n independent repetitions as indicated on the graph. **(B)** Western blot analysis of mutant SERT compared to WT. (Left) Representative immunoblot of total cell lysate derived from transiently transfected HEK293 cells with the specified SERT variant. (Right) Densitometric analysis of immunoblots showed reduced total expression levels of both variants compared to SERT-WT (N=4 independent experiments each normalized to actin and measured as % of SERT-WT). **(C)** Imaging of live HEK293 cells that transiently express WT or mutant SERT following incubation (30 min) with 40 nM of the fluorophore-conjugated antagonist, VK02-83, to label SERT in the plasma membrane. (Left) Bar graph showing the mean VK02-83 membrane intensities of images recorded from three independent experiments normalized to the average of SERT-WT for each experiment. SERT-N217S displayed reduced VK02-83 labelling intensity, indicating impaired expression of functional SERT expressed on the surface, while labelling intensity of SERT-A500T was comparable to SERT-WT. (Right) Representative images of VK02-83 stainings. The scale bar is 10 µM. **(D+E)** (Left) Bar graphs of the co-localization quantified as the Pearson coefficient of WT and mutant SERT with the ER marker BiP **(D)** and Golgi marker Giantin (**E)**. SERT-N217S, but not SERT-A500T, show more colocalization with BiP and Giantin, consistent with impaired trafficking and ER and Golgi retention. The data are from three independent transfections, where the number of images (n) for each SERT variant is indicated on the graph. (Right) Representative images, with scale bar of 10 µM. Symbols: #, P < 0.05; ##, P ≤ 0.01; ###, P ≤ 0.001; ####, P < 0.0001 by one-sample t-test and *, P < 0.05; ***, P ≤ 0.001 by one-way ANOVA with Dunnett’s post hoc analysis comparing each mutant SERT with WT-SERT.

Since the mutations were found in treatment-resistant patients, we next investigated if the disease variants had altered pharmacological profiles towards classical blockers of SERT. To address this, we carried out [^3^H]-5-HT uptake inhibition assays with the SERT blockers *S*-citalopram, venlafaxine, amitriptyline and imipramine (Supplementary Figure 1A-D) to derive inhibition constants (*K_i_*) (summarized in Table 2). For SERT-N217S, we did not observe any changes in inhibitor binding compared to SERT-WT. SERT-A500T showed a small decrease in apparent affinity for amitriptyline (*P* = 0.034, Table 2), but did not demonstrate altered interaction with *S*-citalopram, venlafaxine, or imipramine. Collectively, these data suggest that treatment resistance in the patient carriers of SERT-N217S and SERT-A500T does not arise from impaired affinity to antidepressant agents.

**Table 2.** Pharmacological properties of SERT-N217S and SERT-A500T. p*K*_i_ values for denoted SERT ligands were derived from [^3^H]5-HT uptake competitive inhibition assays on transiently transfected HEK293 cells. All experiments were performed in triplicates. The statistics were performed using one-way ANOVA with Dunnett’s multiple comparisons test.

| Compound | SERT-WT |  |  | SERT-N217S |  |  |  | SERT-A500T |  |  |  |
| --- | --- | --- | --- | --- | --- | --- | --- | --- | --- | --- | --- |
| | $pK_i$ | ±SEM | N | $pK_i$ | ±SEM | N | P | $pK_i$ | ±SEM | N | P |
| S-Citalopram | 8.28 | 0.13 | 8 | 8.25 | 0.03 | 4 | 0.988 | 8.48 | 0.25 | 4 | 0.599 |
| Venlafaxine | 7.41 | 0.05 | 9 | 7.59 | 0.10 | 5 | 0.130 | 7.42 | 0.01 | 3 | 0.995 |
| Amitriptyline | 7.30 | 0.05 | 8 | 7.47 | 0.03 | 4 | 0.160 | 7.05 | 0.11 | 4 | 0.034 |
| Imipramine | 7.94 | 0.12 | 6 | 8.11 | 0.24 | 4 | 0.689 | 7.39 | 0.11 | 3 | 0.088 |
| VK02-83 | 7.57 | 0.12 | 10 | 8.02 | 0.21 | 6 | 0.072 | 7.41 | 0.08 | 5 | 0.694 |

### SERT-N217S shows trafficking impairments and reduced surface expression

To study if the compromised uptake capacities of SERT-N217S and SERT-A500T were caused by altered cellular processing of the disease-associated variants of the transporter, we carried out Western blotting of lysates from transiently transfected HEK293 cells. Both SERT-WT and SERT-A500T transfected cells gave rise to two distinct bands of ∼75 kDa and ∼55-60 kDa, presumably reflecting mature and immature glycosylated SERT (40). The 55 kDa band was also observed for SERT-N217S expressing cells, but the larger band was narrower and migrated at ∼70 kD instead (Figure 2B), consistent with lack of glycosylation at position 217. Importantly, quantification of the total immunosignal showed a marked reduction for SERT-N217S (46.9 ± 7.1% of WT, *P* = 0.0050, Figure 2B), suggesting overall reduced expression of the mutant transporter. The signal for SERT-A500T was also reduced compared to SERT-WT (85.5 ± 3.0% of WT, *P* = 0.014 Figure 2B) suggesting that the SERT-A500T mutation also interferes with the expression of SERT, although less than the SERT-N217S mutation. To confirm the missing glycosylation site caused the changed migration pattern of SERT-N217S, lysates from both SERT-WT and SERT-N217S expressing cells were treated with peptide N-glycosidase (PNGase), which cleaves asparagine-bound N-glycans. Indeed, following the treatment, we observed only the lower band (50-60 kD) on the blot for both WT and SERT-N217S (Supplementary Figure 2), supporting the conclusion that the higher molecular weight bands reflect transporter with different degrees of N-linked glycosylation.

To assess if the SERT-N217S and SERT-A500T mutations alter the expression level of SERT at the cell surface, we used the recently developed fluorescent *S*-citalopram analogue, VK02-83 that enables live visualization of SERT in the plasma membrane (39). Of note, VK02-83 showed similar binding properties at SERT-WT, SERT-N217S, and SERT-A500T (Table 2 and Supplementary Figure 1E). Following live labeling with 40 nM VK02-83, confocal images were acquired of live HEK293 cells expressing SERT-WT, SERT-N217S or SERT-A500T. The images showed a clear membrane labeling for WT and mutant SERT expressing cells (Figure 2C). Compared to SERT-WT, however, the mean fluorescence intensity was significantly lower for SERT-N217S (SERT-WT = 100 ± 7.6% vs. SERT-N217S = 62.5 ± 5.0%, *P* = 0.0002), supporting the conclusion that reduced surface expression accounts for at least a considerable part of the impaired uptake capacity of SERT-N217S. SERT-A500T expressing cells, on the other hand, did not show lower VK02-83 labelling intensity (89.1 ± 5.8%, *P* = 0.37). Importantly, the VK02-83 staining was blocked when pre-and co-incubating with the selective SERT inhibitor, paroxetine (1 µM), confirming that the signal was due to specific SERT binding (Supplementary Figure 3).

As N-glycosylation plays a pivotal role in the protein folding process in the ER and early secretory pathway (52), we next investigated if the altered glycosylation of the SERT-N217S variant causes ER and/or Golgi retention. We used confocal fluorescence-microscopy to conduct co-localization analysis using an antibody against SERT together with markers for ER (BiP/HSP70) (Figure 2D) or Golgi (giantin) (Figure 2E). We used Pearson’s correlation coefficient to quantify the relative degree of co-localization for SERT-WT, SERT-N217S, and SERT-A500T. For SERT-N217S, we found a higher degree of co-localization with both BiP/HSP70 and giantin than for SERT-WT (Figure 2D-E), further supporting SERT-N217S trafficking to the surface is impaired in consequence of disrupted glycosylation. Consistent with the VK02-83 staining, we did not find evidence for ER or Golgi-retention of the SERT-A500T variant, which displayed comparable Pearsons’ coefficients as SERT-WT for both markers. Of notice, we also evaluated the degree of colocalization with the lysosomal marker, Lamp-1, and found no difference between the Pearson’s coefficients for SERT-WT and the two mutants (Supplementary Figure 4).

### SERT-N217S has a dominant-negative effect on 5-HT transport

In addition to directly impairing SERT function, a potentially critical factor for the pathological impact of the hypomorphic disease variants would be their ability to act in a dominant-negative manner to influence the function of SERT-WT. SERT has been shown to assemble into oligomers before and after ER export (53), allowing mutants to compromise the trafficking and/or function of co-expressed SERT-WT. We rationalized that if the mutants have dominant-negative effects, they should adversely affect the uptake capacity of SERT-WT when co-expressed, i.e., the presence of the mutant should reduce SERT-WT activity. To test this, we co-expressed equal amounts (2μg) of SERT-WT and either SERT-N217S or SERT-A500T in HEK293 cells and compared the resulting [^3^H]5-HT uptake capacity with that obtained from cells expressing only SERT-WT together with 2 µg empty vector to ensure that the total amount of DNA was kept constant (4 μg) for all conditions (Figure 3). Most notably, we found that cells expressing the SERT-N217S variant ‘on top’ of SERT-WT displayed significantly lower uptake capacity than SERT-WT only expressing cells (86.5 ± 2.9% of WT, *P* = 0.0033) even though the total amount DNA encoding the transporter was twice as high (2μg SERT-WT + 2μg SERT-N217S). We observed a similar, but non-significant, trend for the SERT-A500T (90.7 ± 3.4% of WT, *P* = 0.053). Importantly, the dominant-negative effect of SERT-N217S was not a result of an unspecific saturating of the translation, maturation or trafficking machinery, as transfection with higher amounts of SERT-WT (4 µg) produced an increase in uptake capacity (116.9 ± 4.6% of 2 µg WT, *P* = 0.0076).

**Figure 3.**
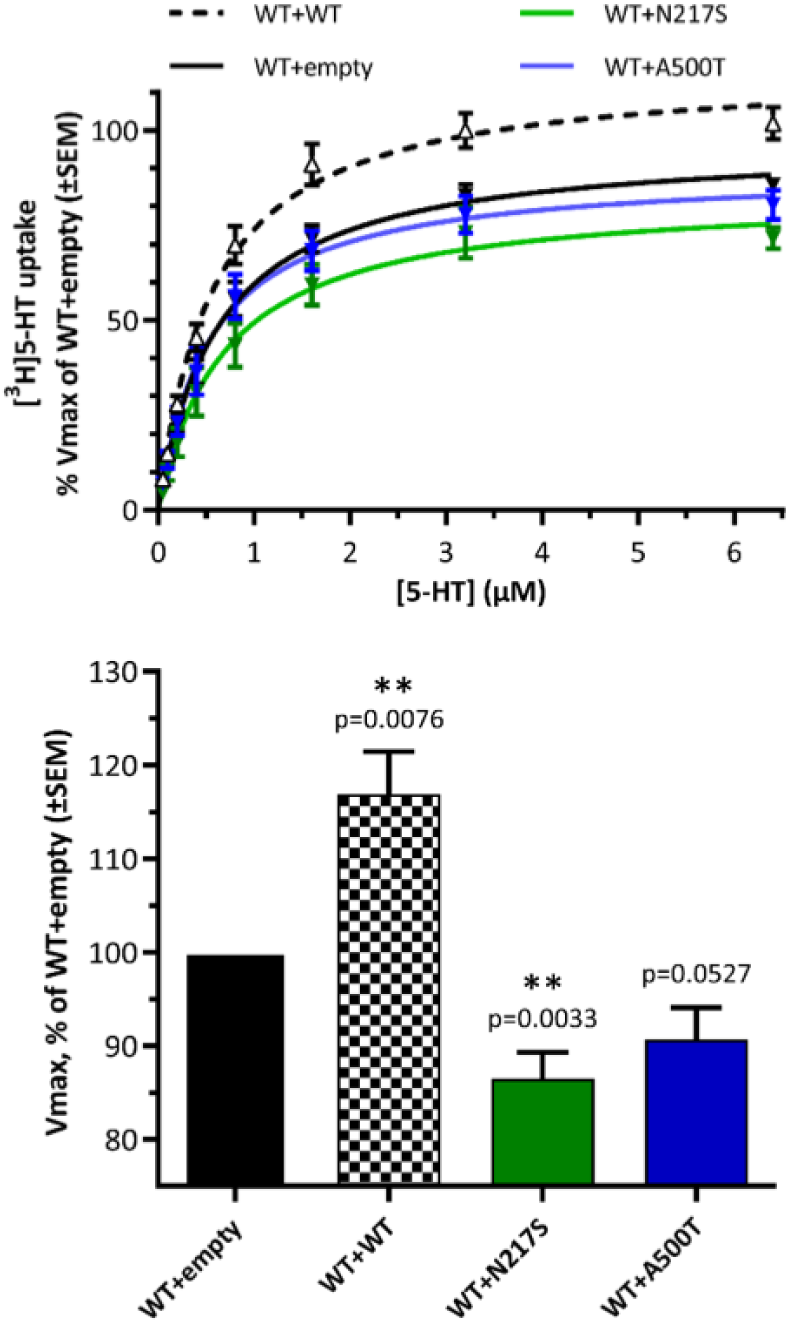
SERT-N217S exhibits dominant-negative effect on SERT-WT function. Dominant negative effects of SERT-N217S and SERT-A500T on SERT-WT were evaluated by comparing the uptake capacity of SERT-WT alone and when SERT mutants were expressed on top of SERT-WT. **(A)** Average [^3^H]5-HT uptake curves from HEK293 cells transiently transfected with 2 µg DNA of SERT-WT and 2 µg DNA of either empty vector (control), SERT-N217S, SERT-A500T or additional SERT-WT as an additional control. **(B)** Bar graph of mean *V*_max_ values in % of WT+empty. Expression of SERT-N217S on top of SERT-WT exerts a dominant negative effect on *V*_max_. Data are mean ± SEM from 5-12 experiments, each performed in triplicates and normalized to the *V*_max_ of WT+empty cells. (N(WT+empty)=12, N(WT+WT)=8, N(WT+N217S)=7 and N(WT+A500T)=5, one-sample t-test, with Bonferroni adjustments of significance level to 0.0167). Symbols: **, P ≤ 0.01.

### Pharmacochaperoning rescues the SERT-N217S variant

Our functional investigations of SERT-N217S and SERT-A500T uncovered that both variants are hypomorphic, suggesting that the loss of SERT activity could be central to the disease association. Therefore, we evaluated if the variants are amenable for pharmacochaperoning as a means to increase surface expression and function (54, 55). It has been shown that noribogaine stabilizes SERT in an inward-open conformation and that this property can help rescue engineered folding-deficient SERT mutants to pass through the cell’s quality-control system (56). Furthermore, pharmacological chaperoning has been shown to rescue clinically relevant hypomorphic dopamine transporter (DAT) variants linked to DAT deficiency syndrome *in vitro* and *in vivo* (57–59). Thus, we assessed if we could rescue the disease-associated SERT variants by pharmacological chaperoning with noribogaine. We transiently transfected HEK293 cells with SERT-WT, SERT-N217S or SERT-A500T and incubated the cells with or without noribogaine (10 µM) for 24 h before measuring total protein expression and surface expression by surface biotinylation and western blotting (Figure 4). For SERT-N217S, we found that noribogaine increased both total protein expression (160.3 ± 10.5% of vehicle *P* = 0.011) and surface expression (157.6 ± 11.0% of vehicle *P* = 0.013). A similar non-significant trend was observed for SERT-A500T (total expression = 136.3 ± 16.6% of vehicle, *P* = 0.12 and surface expression = 140.1 ± 17.7% of vehicle, *P* = 0.11). In contrast, noribogaine did not produce robust changes in SERT-WT total protein expression (105.9 ± 5.3% of vehicle *P* = 0.35) or surface expression (108.8 ± 15.4% of vehicle, *P* = 0.61, Figure 4). These findings demonstrate the feasibility of using conformationally stabilizing agents to remedy the potential deficits of trafficking impaired loss-of-function variants.

**Figure 4.**
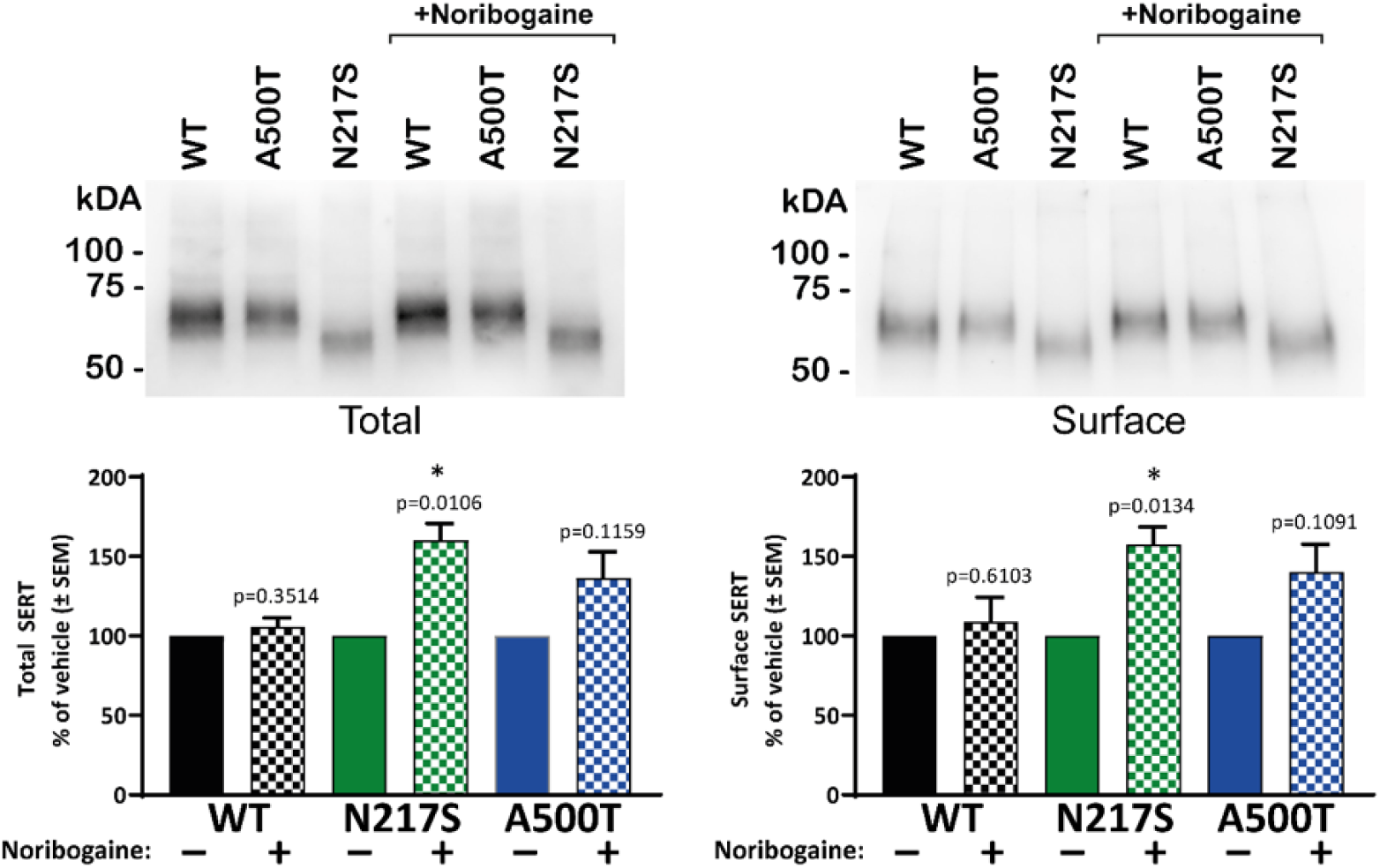
Expression SERT-N217S is amenable to pharmacochaperoning. Analysis of total and surface protein expression by biotinylation following treatment with the potential pharmacochaperone, noribogaine. Cultures of HEK293 cells transiently expressing either SERT-WT, SERT-N217S, or SERT-A500T were treated with vehicle or 10 µM noribogaine for 24 h before assaying. Immunoblotting was used to detect either whole-cell SERT expression (left) or only surface biotinylated SERT (right). (Top) representative immunoblots. (Bottom) Densitometric analysis of immunoblots with each genotype normalized to its own vehicle control in each experiment. Both total and surface expression of SERT-N217S was increased by the noribogaine treatment while SERT-A500T and SERT-WT did not show significant changes (N=4, one-sample t-test, with Bonferroni adjustments of significance level to 0.0167). Data are mean ± SEM. Symbols:*, P < 0.05.

## DISCUSSION

Being a key regulator of 5-HT neurotransmission, SERT stands out as the only monoamine transporter for which no loss-of-function missense mutation has been linked to diseased states (37, 53, 60–62). Thus, while SERT gain-of-function coding variants have been linked to OCD and ASD, the consequences of hypomorphic coding SERT variants remains unknown (27–30).

Here we investigate rare genetic variants in SLC6A4 in a sample of patients with affective disorders, uniquely selected for a highly treatment-resistant disease phenotype, reflected in a lifetime history of ECT treatment. The rationale for specifically investigating the occurrence of coding SERT variants in this sample is twofold. First, SERT is a principal target for antidepressant medications, and coding variants of the transporter could have direct pharmacogenetic implications on treatment responses (28, 63). Second, serotonergic disturbances have long been hypothesized to participate directly in the etiology of affective disorders (17, 64–66). Given the critical role of SERT controlling extracellular 5-HT and maintaining intracellular stores, altered SERT function could directly inflict disease-relevant changes in serotonergic circuits.

Among 144 patients, we identified three unrelated patients that were heterozygote for two coding SERT variants that have not been described or characterized previously. Two patients carried the same SERT-N217S variant and the third carried the SERT-A500T variant. Moreover, the two patients carrying the SERT-N217S mutation both had a family history of neuropsychiatric disease with several close relatives suffering from mental disorders related to 5-HT dysfunction, including affective disorders, anxiety, schizophrenia and alcohol use disorder. We did not find a similar striking accumulation of psychiatric diagnoses among close relatives to the patient carrying the SERT-A500T mutation. Unfortunately, without genotype information from the relative, we cannot tell if SERT-A500T is a de novo variant. Even with such information, the available pedigrees for all three patients would still be too small for proper linkage analysis. Instead, by comparing the allele frequencies in our cohort with the frequencies found in the gnomAD database (45) and in an ethnicity-matched control population from the iPSYCH cohort (41), we determine that both SERT variants accumulate significantly in our cohort, supporting that SERT-N217S and SERT-A500T are pathogenic risk variants for affective disorder.

The genetic disease association is further strengthened by our functional studies of SERT-N217S and SERT-A500T, revealing important insights into how the mutations affect SERT function. Most importantly, we found that SERT-N217S and SERT-A500T are partial loss-of-function variants with ∼46% and ∼63% residual uptake capacity compared to SERT-WT mediated 5-HT uptake *in vitro*. To our knowledge, the only previously characterized missense loss-of-function SERT coding variant, identified in a human population, is SERT-P339L. However, this variant was found in a sample of anonymous individuals without available information on clinical phenotypes (28, 67). Thus, identifying two different functionally impaired coding SERT variants enriched in a cohort of patients with treatment-resistant affective disorders is striking.

While few studies have addressed the role of rare coding SERT variants in psychiatric diseases, common variants have been widely investigated in early candidate gene studies. The most extensively researched common polymorphism of SERT is the non-coding variable number of tandem repeat polymorphism, 5-HTTLPR, located in the promoter region of *SLC6A4.* It exists as a long ‘l’ allele, linked to high SERT expression, and a short ‘s’ allele, linked to low SERT expression (68). Interestingly, meta-analyses have reported an association between the low expressing ‘s’ 5-HTTLPR allele and major depressive disorder (69, 70). The ‘s’ allele has also been linked to anxiety-related personality traits (71) and studies on gene-environment interactions have reported that individuals with the ‘s’ allele of 5-HTTLPR exhibit more depressive symptoms as a function of stressful life events (70), which is interesting as we found similar phenotypes both among the SERT-N217S carriers and among their family members. Finally, the impaired remission and slower response rate to SSRIs has been reported for depressive patients that are homozygous for the ‘s’ 5-HTTLPR allele (72), which is again consistent with the treatment-resistant classification of the index patients we describe. However, the disease associations to 5-HTTLPR have not been consolidated by GWAS studies (10–13). Our investigations of the rare coding SERT-N217S and SERT-A500T variants with potentially larger effect size, therefore, provide important new support to the longstanding hypothesis that disturbances in 5-HT signaling arising from SERT dysfunction confer risk for affective disorder.

Our in vitro characterization demonstrated a shared molecular phenotype for SERT-N217S and SERT-A500T manifesting as impaired 5-HT uptake capacity but also revealed that the two mutations impair SERT function through different mechanisms. The SERT-N217S mutation directly abolishes one of the two glycosylation sites present in the second extracellular loop of SERT. Despite being predicted to be a benign variant, our investigations revealed a pronounced loss of uptake capacity by SERT-N217S. This impaired function does not reflect disrupted substrate binding, as we found that the SERT-N217S variant even has slightly increased apparent affinity for 5-HT. Instead, western blot analysis and visualization of SERT-N217S at the cell surface by using a *S*-citalopram-based fluorescent ligand (39) showed that SERT-N217S displays reduced expression and surface targeting. Glycosylation is known to be essential for proper folding and trafficking through the secretory pathway (52), and we observed increased co-localization between SERT-N217S and markers for ER and Golgi compared to SERT-WT, supporting that the expression of SERT-N217S is compromised, at least in part by incomplete or slower folding and trafficking. Further supporting that the retention of the SERT-N217S variant results from a folding deficiency, we observed that molecular chaperoning with noribogaine increased both surface and total protein expression of SERT-N217S but not of SERT-WT.

Our studies on SERT-A500T confirmed the predicted deleterious consequences of the mutation as it displays a residual uptake capacity of ∼63% of SERT-WT. While we observed a slight decrease in total protein expression of SERT-A500T, compared to WT, we did not find significantly reduced surface expression when imaging surface labeling with VK02-83, suggesting that the impaired uptake by SERT-A500T arises primarily from impaired catalytic activity rather than impairments in surface targeting. Consistently we did not find alterations in the colocalization of SERT-A500T with markers of ER and Golgi, and we only observed a trend in the rescue of surface expression of SERT-A500T by noribogaine. The SERT-A500T mutation is located in the center of transmembrane helix 10 in a tight space between sidechains in TM3 and TM12. Therefore, a mutation from alanine to threonine could potentially destabilize the transmembrane region since this specific site has limited space to accommodate the bulkier sidechain of threonine. Furthermore, threonine is a β-branched amino acid which is disfavored in alpha-helices as it adds a hydroxyl group in the hydrophobic transmembrane domain (73), which again could compromise the catalytic function of the transporter.

An interesting question that arises from our finding is how impaired SERT function might sensitize a person to develop an affective disorder. SERT-mediated reuptake is critical for regulating quantal size and the spatiotemporal propagation of 5-HT signals. Compromised SERT function is therefore likely to produce marked changes in serotonergic neurotransmission and, accordingly, serotonin-regulated developmental processes and phenotypes. In this respect, another important finding is that at least the SERT-N217S variant displays direct dominant-negative actions on SERT-WT, i.e., the presence of SERT-N217S impairs SERT-WT function. A model to explain how a dominant-negative effect can manifest is that SERT oligomerizes before export from the ER (53, 55). Thus, we propose that SERT-N217S exerts its dominant-negative effect by co-assembly with SERT-WT, thereby hindering the trafficking of both variants through the secretory pathway. In addition, both SERT-A500T and SERT-N217S could affect SERT-WT function when assembled at the plasma membrane in oligomer complexes. Further studies to understand the exact nature of the 5-HT dysfunction that arises from disease-associated hypomorphic SERT variants like SERT-N217S and SERT-A500T *in vivo* are warranted. Still, it seems likely that dominant-negative properties, for instance, can be an essential factor for the effect size of a coding variant.

Finally, our data also suggest that molecular chaperoning could be a possible therapeutic approach to rescue SERT function in patients carrying loss of function coding SERT variants. Intriguingly we noticed that a SERT N208S variant had been reported in the GnomAD database with a considerable allele frequency of 0.13% among Ashkenazi Jews (45). Residue N208, like N217, is an N-glycosylation site that supports SERT surface expression (40). Thus, according to our data on SERT-N217S, the N208S mutation could be another example of a rare coding SERT variant acting as a risk factor for affective disorders with a large effect size.

Collectively, our data expands the spectrum of disease-associated coding SERT variants and suggests that genetic insults to SERT function can drive neural changes that are sensitizing towards affective disorders and anxiety.

## Data Availability

The data underlying this study are not publicly available due to the Danish Data Protection Act and European Regulation 2016/679 of the European Parliament and of the Council (GDPR) that prohibit distribution of personal data. The data are available from the corresponding authors upon reasonable request and under a data transfer and collaboration agreement.

## ACKNOWLEDGEMENTS

We wish to thank Anette Dencker Kaas for her outstanding technical assistance, Troels Rahbek-Clemmensen for assisting sanger sequencing analysis, and Niels Felsted for helping out with data managemen.t We thank the iPSYCH PIs Professors Merete Nordentoft, Preben Bo Mortensen, Anders Børglum, Ole Mors, and Dr. David Hougaard along with Drs. Vivek Appadurai and Alfonso Demur for look-up in the iPSYCH database. This work was supported by: Independent Research Fund Denmark – Medical Sciences (DFF-4183-00571, FH; DFF-4004-00097B, UG), Lundbeck Foundation (R181-2014-3090 and R303-2018-3540, FH), Lundbeck Foundation (R223-2016-261, UG), National Institute on Drug Abuse-Intramural Research Program (Z1A DA000610, AHN), and NIH award MH094527 (RDB).

## CONFLICT OF INTEREST

All authors report no biomedical financial interests or personal relationships that could have influenced the work written in this paper.

Supplementary information is available at MP’s website.

