## Supplementary information for "Disruptive mutations in the serotonin transporter associate serotonin dysfunction with treatment-resistant affective disorder"

^1^Molecular Neuropharmacology and Genetics Laboratory, Department of Neuroscience, Faculty of Health and Medical Sciences, University of Copenhagen, Denmark. ^2^Institute of Biological Psychiatry, Mental Health Services Copenhagen; Department of Clinical Medicine, University of Copenhagen; and The Lundbeck Foundation Initiative for Integrative Psychiatric Research (iPSYCH), Denmark. *On behalf of iPSYCH researchers. ^3^Medicinal Chemistry Section, National Institute on Drug Abuse-Intramural Research Program, National Institutes of Health, Baltimore, USA. ^4^Stiles-Nicholson Brain Institute and Department of Biomedical Science, Charles E. Schmidt College of Medicine, Florida Atlantic University, Jupiter, Florida, USA.

† Address author correspondence to: Freja Herborg, Department of Neuroscience, Maersk Tower 7.5, University of Copenhagen, Blegdamsvej 3B, DK-2200 N, Copenhagen, Denmark. Phone +4553609699;.

SUPPLEMENTARY FIGURES

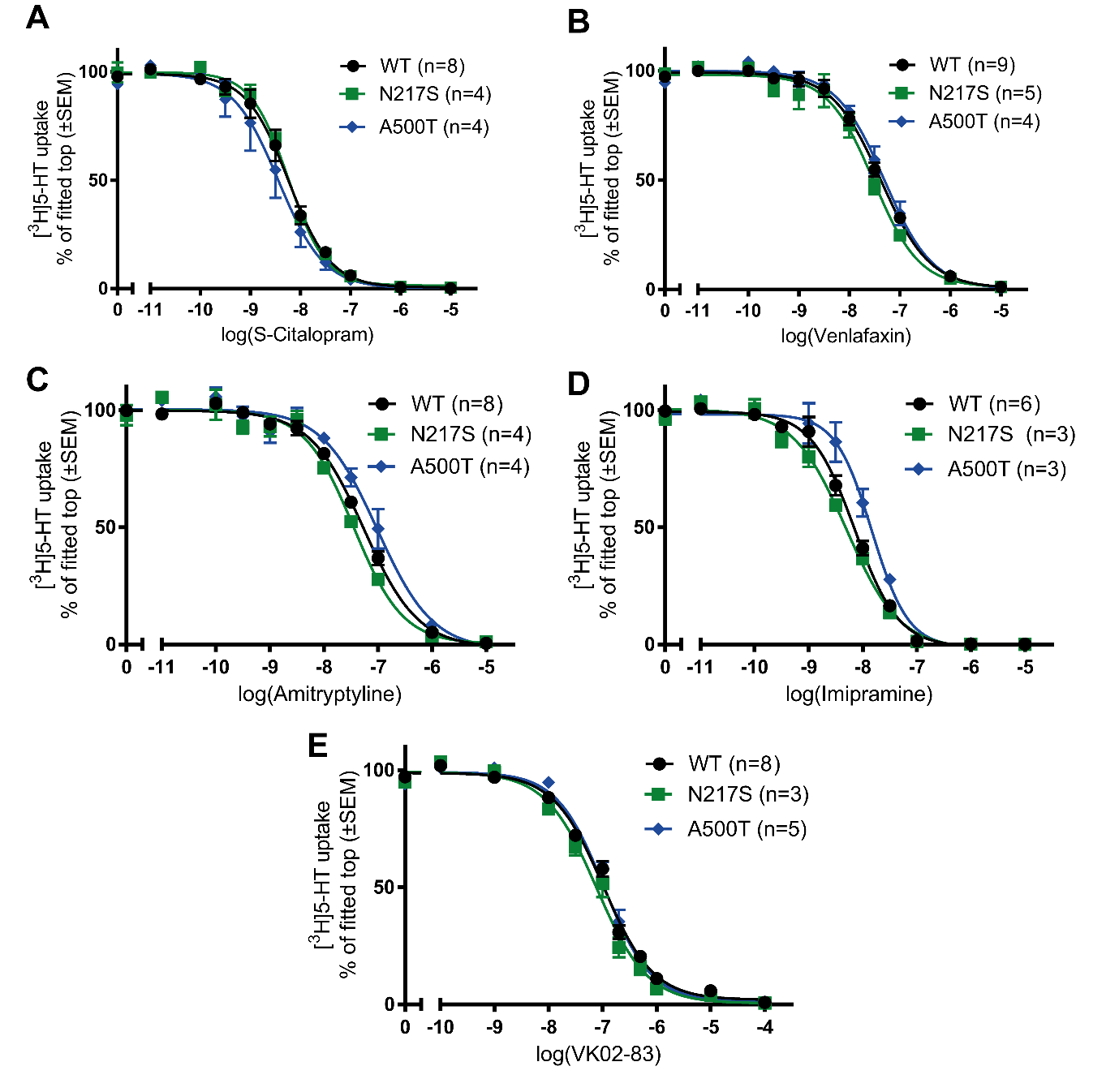

**Supplementary figure 1** - Pharmacological profile of WT SERT and the two mutants N217S and A500T. A-E) Competition-binding assay with A) *S*-Citalopram, B) Venlafaxine, C) Amitriptyline, D) Imipramine and E) VK02-83. All experiments were performed in triplicates with n repetitions as stated in the each graph.

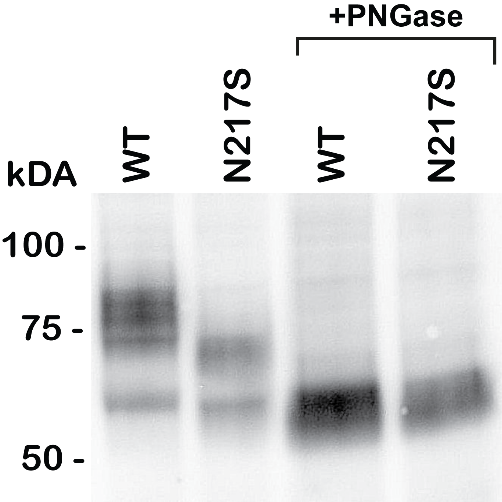

**Supplementary figure 2** - Immunoblotting of total cell lysates derived from HEK293 cells transiently expressing WT SERT or the N217S variant with and without PNGase treatment before blotting demonstrates the missing upper molecular weight band of SERT-N217S reflects a difference in glycosylation. The blot is representative of 4 independent experiments.

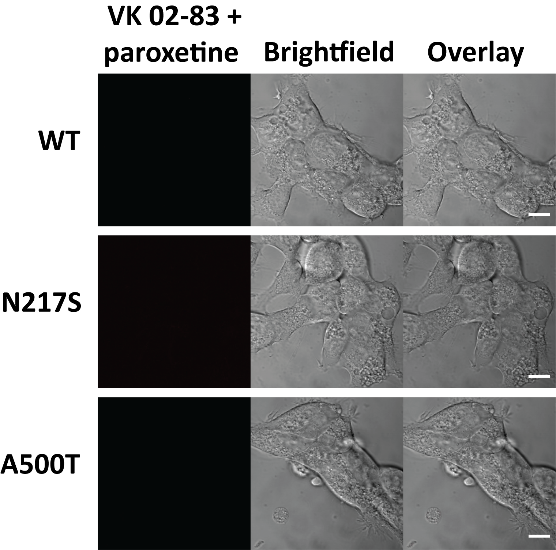

**Supplementary figure 3** - VK02-83 causes specific staining of mutant and WT SERT as no VK02-83 staining is seen following pre- and co-incubation with 1 µM paroxetine. Representative images of VK02-83 staining from three independent experiments. The scale bar is 10 µm.

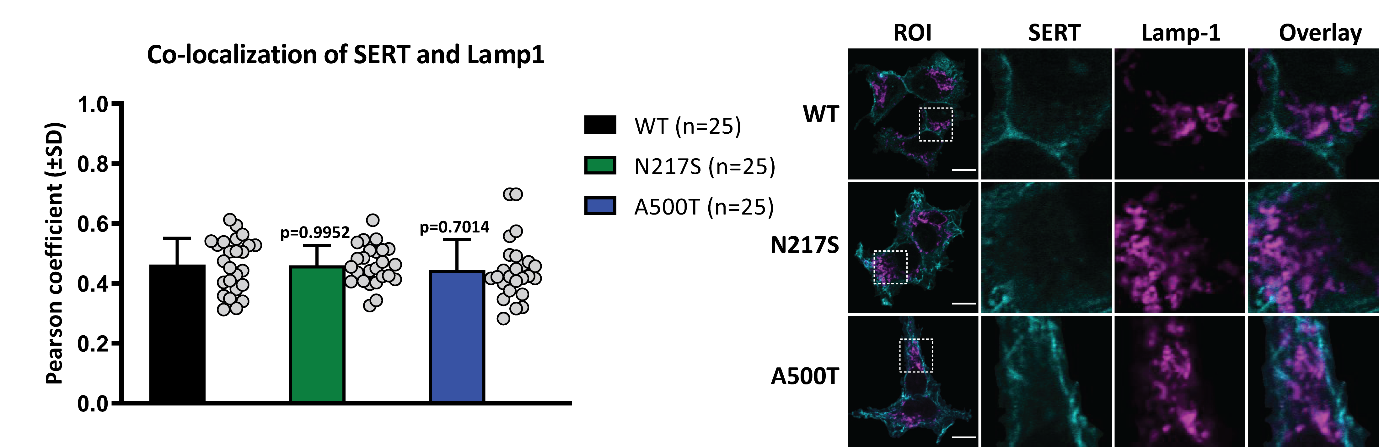

**Supplementary figure 4** - No increase in lysosomal localization was seen for SERT-N217S and SERT-A500T compared to WT-SERT. (Left) Bar graphs of the co-localization quantified as the Pearson coefficient of WT or mutant SERT and the lysosomal marker Lamp1. Data is based on three independent experiments resulting in a total of n number of images per condition as stated in right-hand side of the graph. (Right) Representative images, in which the scale bar is 10 µm. One-way ANOVA with Dunnett’s post hoc analysis compared SERT-N217S and SERT-A500T with WT-SERT.

SUPPLEMENTARY METHODS

**[^3^H]-5-HT Uptake**

Uptake experiments were performed on transiently transfected HEK293 cells, expressing WT SERT, SERT-N217S, or SERT-A500T as described previously (1). Briefly, cells were seeded in 24 well plates (150.000 cells per well) coated with polyornithine 24 h prior to the experiments. Uptake experiments were carried out using [^3^H]-5-HT diluted 1:200 to ~10nM in uptake buffer (25 mM HEPES, 120 mM NaCl, 5mM KCl, 1.2 mM CaCl_2_, 1.2 mM MgSO_4_, 1 mM ascorbic acid, 0.1 mM pargyline, 5 mM D-glucose and pH adjusted to 7.4 with NaOH). The presence of 100 µM paroxetine was used to determine non-specific 5-HT binding. For saturation kinetic measurements, [^3^H]5-HT was mixed with unlabeled 5-HT in a 2-fold dilution row from 6.4-0.05 µM in uptake buffer with a constant specific activity. *K*_m_ and *V*_max_ values were obtained from fitting each experiment to the Michaelis-Menten equation in GraphPad Prism. 5-HT and SERT inhibitor affinities were determined using a fixed concentration of [^3^H]5-HT and increasing concentrations of the specified compound. Estimation of *K*_i_ was calculated using the equation *K*_i_ = IC_50_/(1+(L/*K*_m_)) where L is the concentration of [^3^H]5-HT and the IC_50_ were estimated by fitting each experiment to a sigmoidal dose-response curve in GraphPad Prism (version 9.3.1).

Imaging Analysis

For membrane intensity quantification, the image was converted to 8-bit before running a median filter. Thresholding was done using ImageJ’s build-in Li’s Minimum Cross-Entropy thresholding method and the resulting mask was applied on the original unmodified image to measure the average membrane intensity. For the co-localization assay, images were first cropped to exclude cells only partially in-frame. The build-in Huang’s fuzzy thresholding method was used on the channel containing the 488nm (SERT) signal. From this, ROIs containing whole cells were included and the rest was cropped away from both channels (488 and 543nm) of each image. The Pearson coefficient was then calculated on the resulting image using the ImageJ plugin JACoP (2, 3).

**Surface Biotinylation**

Briefly, transiently transfected cells were washed with ice-cold PBS, incubated in cold 1 mg/mL sulfo-NHS-SS-biotin in PBS for 40 min on ice, and washed twice with 100 mM glycine in PBS following two additional washes in PBS. Cells were then lysed in solubilization buffer (25 mM Tris, pH 7.5 with 150 mM NaCl, 1% Triton X-100, 0.2 mM PMSF and complete protease inhibitor cocktail from Roche) and incubated for 20 min with end-over-end rotation at 4°C centrifuged for 20 min at 16,000 g. The protein content was quantified with a BCA Protein Assay Kit (Pierce Biotechnology) and was distributed with equal protein concentration on avidin beads and incubated overnight at 4°C. Following centrifugation, the pellets containing the avidin beads were washed 4 times with solubilization buffer before elution with 2x loading buffer (100 mM Tris-HCl, pH 6.8 containing 100 mM DTT, 20% glycerol, 10% SDS and 0.2% bromophenol blue) for 30 min at 37°C. Total lysates were equally incubated in a 2x loading buffer for 30 min. Following elution, the avidin beads were excluded by filtration. The biotinylated and total lysate samples were separated by SDS-PAGE and SERT was detected by Western blotting.

**Western Blotting**

Lysates were prepared and protein concentration determined as described for surface biotinylation Following protein concentration determination, samples were diluted in 4x SDS loading buffer and incubated at 37°C for 30 min. SDS-PAGE with Any kD Gels (Bio-Rad) was used to separate proteins following transfer to Immobilon-p membranes (Millipore). Blocking of membranes was done by incubation for 1 h in blocking buffer (0.05% Tween-20 and 5% dry milk in PBS) and membranes were then incubated in blocking buffer with primary goat anti-SERT antibody (Santa Cruz Biotechnology, sc-1458; discontinued, 1:750) for 1h at RT or 4°C overnight. This was followed by incubating the membrane with secondary horseradish peroxidase-coupled rabbit anti-goat antibody (Pierce, 1:5000) for 1h at RT. The blots were washed and visualized by ECL kit (Amersham Biosciences) for chemiluminescence with AlphaEase (Alpha Innotech). For loading control, the membranes were re-probed against β-actin (Sigma-Aldrich). Densitometric analysis of bands was done in ImageJ.

ImageJ Macros

VK02-83 membrane quantification macro:

dir=getDirectory("Choose a Directory");

list = getFileList(dir);

run("Set Measurements...", "area mean min limit display redirect=None decimal=3");

for (i=0; i<list.length; i++) {

open(dir+list[i]);

selectWindow(list[i]);

run("Duplicate...", "title=copy");

setOption("ScaleConversions", true);

run("8-bit");

run("Median...", "radius=3 slice");

run("Auto Threshold", "method=Li ignore_black white");

run("Convert to Mask");

run("Divide...", "value=255.000 slice");

open(dir+list[i]);

run("8-bit");

imageCalculator("Multiply create 32-bit", list[i], "copy");

selectWindow("Result of "+list[i]);

//run("Threshold...");

setThreshold(1, 10000000000);

run("Measure");

run("Close All");

}

Crop image Macro:

dir=getDirectory("Choose a Directory");

newDir = substring(dir,0,lengthOf(dir)-1) + " cropped";

if (File.isDirectory(newDir) == false) {

File.makeDirectory(newDir);

}

list = getFileList(dir);

for (i=0; i<list.length; i++) {

if (endsWith(list[i], ".lsm")) {

run("Clear Results");

if (roiManager("count")>0) {

roiManager("reset");

}

name = list[i];

path = dir + name;

newname = substring(name, 0, lastIndexOf(name, ".lsm"));

open(path);

run("Median...", "radius=6 stack");

run("Auto Threshold", "method=Huang ignore_black white");

run("Analyze Particles...", "size=50-Infinity add stack");

if (roiManager("count")!=0) {

close();

open(path);

run("Original Scale");

roiManager("show all with labels");

beep();

Dialog.create("Select ROIs");

for (j=0; j<roiManager("count"); j++) {

Dialog.addCheckbox(j+1, false);

}

Dialog.show();

rois=newArray();

for (k=0; k<roiManager("count"); k++) {

if (Dialog.getCheckbox()) {

rois=Array.concat(rois, k);

}

}

if (lengthOf(rois)>0) {

roiManager("deselect");

roiManager("select", rois);

if (lengthOf(rois)>1) {

roiManager("combine");

}

run("Clear Outside", "stack");

roiManager("reset");

selectWindow(name);

saveAs("Tiff", newDir+"/"+newname+".tif");

run("Close All");

} else {

print("No ROIs selected for "+list[i]);

run("Close All");

}

} else {

print(list[i]+" didnt have any ROIs");

run("Close All");

}

} else {

print(list[i]+" is not a lsm file");

}

}
